# Effectiveness of Hyperbaric Oxygen Therapy Compared to Surgical Free Tissue Flap for Osteoradionecrosis of the Jaw: A Meta-Analysis

**DOI:** 10.1101/2023.09.25.23296038

**Authors:** Megha K. Sheth, Casey Collet, De-Chen Lin, Uttam K. Sinha

**Affiliations:** Department of Otolaryngology, Keck School of Medicine, University of Southern California, Los Angeles, CA 90033, USA; Center for Craniofacial Molecular Biology, Herman Ostrow School of Dentistry, and Norris Comprehensive Cancer Center, University of Southern California, Los Angeles, CA 90033, USA

## Abstract

**Background:** Osteoradionecrosis (ORN) of the jaw is an uncommon but potentially serious complication of radiation therapy for head and neck cancer, with many cases presenting months to years after completion of radiotherapy. ORN-related morbidity is high, making effective management of osteoradionecrosis essential. Current treatment options include hyperbaric oxygen therapy (HBOT), surgical management (free flap), and PENTOCLO, among others. Our goal is to analyze the reported efficacy of hyperbaric oxygen therapy versus microvascular free flap in ORN.

**Methods:** This review was conducted using the Preferred Reporting Items for Systematic Review & Meta-Analysis (PRISMA) guidelines. A systematic review search strategy was developed by a librarian (JED). The search was performed in the following databases on March 8, 2023: PubMed, EMBASE, and ClinicalTrials.gov. Two reviewers independently assessed the titles, abstracts, and full-text manuscripts based on predetermined inclusion and exclusion criteria. The risk of bias of the final included studies was assessed using the Newcastle-Ottawa Scale. final included studies were assessed using the Newcastle-Ottawa risk of bias assessment, and analyses were performed using risk ratios.

**Results:** The initial search yielded 1,614 articles with 407 undergoing full-text review. Ultimately, nine studies met the eligibility criteria and were included in the review. In total, 339 patients were included across the nine studies. Each of the nine studies reported on resolution of ORN with HBOT versus surgical free flap, with six studies showing significant differences in outcomes, with surgical free flap yielding significantly higher complete resolution of ORN compared to HBOT (RR 0.42, 95% CI 0.2-0.85; RR 0.07, 95% CI 0-0.99; RR 0.04, 95% CI 0-0.57; RR 0.12, 95% CI 0.04-0.34; RR 0.21, 95% CI 0.08-0.54; RR 0.43, 95% CI 0.24-0.78). Three studies showed no significant differences in ORN resolution (RR 0.33, 95% CI 0.03-4.19; RR 0.33, 95% CI 0.03-4.19; RR 0.34, 95% CI 0.11-1.09). The summary outcome of ORN resolution found a significant difference between surgical free flap and HBOT (RR 0.28, 95% CI 0.17-0.45).

**Conclusions:** Despite the traditional practice of recommending HBOT for ORN treatment, our meta-analysis suggests that HBOT provides little benefit, especially for later stages of disease, and that surgical intervention via surgical free flap yields superior outcomes. However, there may be some benefit of HBOT in conservative management of early and intermediate cases of ORN. Surgical reconstruction with microvascular free flap may be a superior alternative to HBOT in the management of these patients.

## INTRODUCTION

Osteoradionecrosis (ORN) of the mandible is a devastating complication in head and neck cancer patients treated with radiotherapy. ORN involves areas of exposed devitalized bone in the absence of local neoplastic disease that does not heal within 3 months.^1^

Individuals at risk of developing ORN include those with primary cancer originating in the oral cavity or oropharynx who received radiation doses ≥ 65 Gy and/or underwent post-radiation tooth extractions or implant placements.^1,2^ The reported incidence of ORN varies widely, ranging from 1-30% and typically develops within the first 3 years of radiation therapy (cite this).^2,3^ Incidence has reportedly decreased since the introduction of intensity-modulated radiation therapy (IMRT), as this technology allows the precise delivery of radiation to the tumor while minimizing radiation exposure to the surrounding tissues.^2,4^

Treatment of ORN ranges from conservative management to invasive surgical resection and reconstruction. Conservative management includes antibiotics, analgesics, oral hygiene, tissue debridement, sequestrectomy, hyperbaric oxygen therapy (HBOT), and anti-radiation fibrosis drugs, pentoxifylline and tocopherol.^5^ Surgical management involves more radical resection, often requiring free flap reconstruction.^5^ There is a lack of consensus regarding the role of HBOT in ORN management and therefore the utility of conservative management in staging ORN.

In 1983, Marx et al. introduced the role of HBOT in managing ORN.^6^ He attributed the intervention’s success to its ability to promote neovascularity and neocellularity in chronic hypoxic tissue.^6^ Marx’s protocol has become a widely accepted system as a method of grading ORN. Patients are staged according to their responses to 30 preoperative treatments with HBOT, involving 90-minute dives at 100% oxygen under either 2.0 or 2.4 atmospheres of pressure.^7^ Stage 1 ORN includes exposed alveolar bone in the absence of pathologic fracture, which responds to HBOT and minor bony debridement. Stage 2 disease does not respond to 30 daily HBOT treatments with minor bony debridement, or they require major bony debridement. These patients require more radical surgical debridement plus ten postoperative HBOT treatments. Stage 3 disease involves patients who either failed treatment in stage 1 or 2 or initially presented with pathologic fracture, orocutaneous fistula, or evidence of lytic involvement of inferior mandibular border. These patients require mandibular segmental resection of all necrotic bone in addition to 30 preoperative and 10 postoperative HBOT treatments.

Despite the wide acceptance of Marx’s grading protocol, a consensus on the efficacy of HBOT in ORN management has not been reached, partially attributed to the limited incidence of ORN at a single institution. Therefore, a meta-analysis was conducted comparing treatment outcomes in patients who received HBOT versus those who underwent radical surgical resection and reconstruction.

## METHODS

This review was conducted using the Preferred Reporting Items for Systematic Review & Meta-Analysis (PRISMA) guidelines. The search was performed in the following databases on March 8, 2023: PubMed, EMBASE, and ClinicalTrials.gov. When ClinicalTrials.gov was searched, 3 studies were found. Of these, 2 studies did not meet inclusion criteria and one study was terminated due to poor recruitment. The full search strategy for each database, as well as the inclusion and exclusion criteria used for this study, can be found in the appendix.

All citations (1,615) were imported into EndNote X8.0.1 (Thomson Reuters). The library was exported from EndNote and uploaded into Covidence (Covidence.org) and deduplicated, for a total of 1,104 studies for screening. All study’s titles and abstracts were screened. Full-text manuscripts of potentially eligible articles (407) were independently reviewed by two authors.

Full-text articles were assessed based on the eligibility criteria and then were included for data extraction. The reference list of the full-text articles were hand-searched for identifying additional studies. A third reviewer was not required as disagreements regarding inclusion of the study were resolved by discussion. After review, nine articles were suitable for quantitative synthesis. The Newcastle Ottawa Scale was used to assess risk of bias.

## RESULTS

The initial search yielded 1,614 articles. In total, 510 duplicate records were deleted. We screened 1,104 titles and abstracts and excluded 697 studies which did not meet our inclusion criteria. 407 full-text manuscripts were assessed. Ultimately, nine studies met the criteria for eligibility and were included in the review. **Figure 1** shows the PRISMA flowchart of the search and selection process. **Table 1** summarizes the final included studies. In total, 339 patients were included across all nine studies. Studies from six countries were included. Manuscripts were published between 1983 and 2023. There was a median of 37 patients per study (1-105 patients). There were seven retrospective cohort studies, one case control study, and one case report, reporting on patients with osteoradionecrosis of the jaw, who were treated with HBOT and surgical intervention via free flap.

**Table 1:** Data summary of treatment outcomes with HBOT versus surgical free flap from the nine studies included in the meta-analysis.

|  | Hyperbaric oxygen therapy |  |  | Surgical free flap |  |  |
| --- | --- | --- | --- | --- | --- | --- |
|  | HBOT treated | HBOT resolved | Percentage resolved | Free flap treated | Free flap resolved | Percentage resolved |
| Vanderpuye 2000 | 1 | 0 | 0.0 | 1 | 1 | 100.0 |
| Singh 2023 | 1 | 0 | 0.0 | 1 | 1 | 100.0 |
| Oh 2009 | 10 | 4 | 40.0 | 27 | 27 | 100.0 |
| Jenwitheesuk 2021 | 9 | 0 | 0.0 | 22 | 17 | 77.3 |
| Hahn 1983 | 13 | 0 | 0.0 | 35 | 34 | 97.1 |
| Dieleman 2017 | 27 | 3 | 11.1 | 27 | 26 | 96.3 |
| D'Souza 2014 | 28 | 4 | 14.3 | 28 | 19 | 67.9 |
| D'Souza 2007 | 16 | 3 | 18.8 | 11 | 6 | 0.5 |
| Chen 2016 | 19 | 7 | 36.8 | 86 | 74 | 86.0 |
| Total | 124 | 21 | 16.9 | 238 | 205 | 86.1 |

**Figure 1:**
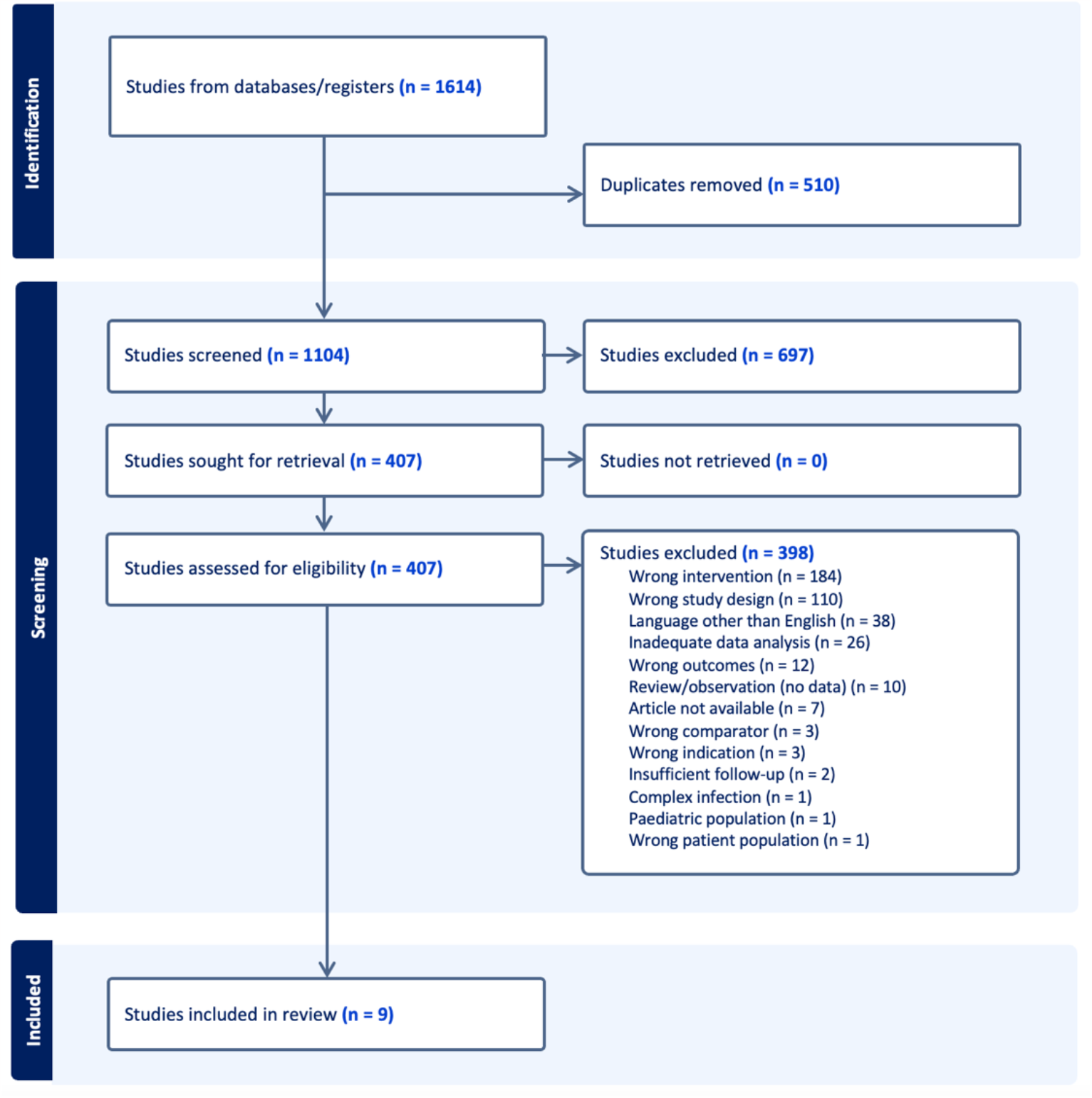
The PRISMA flowchart of the search and selection process for studies analyzing ORN treatment with HBOT versus surgical free flap.

Two studies reported on both clinical improvement and resolution of ORN, while the rest reported only on complete resolution of ORN. The analyses were therefore split into these respective categories. Of the studies reporting on improvement of ORN, one showed significant improvement with surgical intervention compared with HBOT (RR 0.2, 95% CI 0.09-0.42) while the other study showed no significant difference (RR 0.33, 95% CI 0.07-1.51). The summary outcome for ORN improvement was no significant difference between surgical free flap and HBOT (RR 0.22, 95% CI 0.02-2.94). These results are shown in **Table 2**.

**Table 2:**
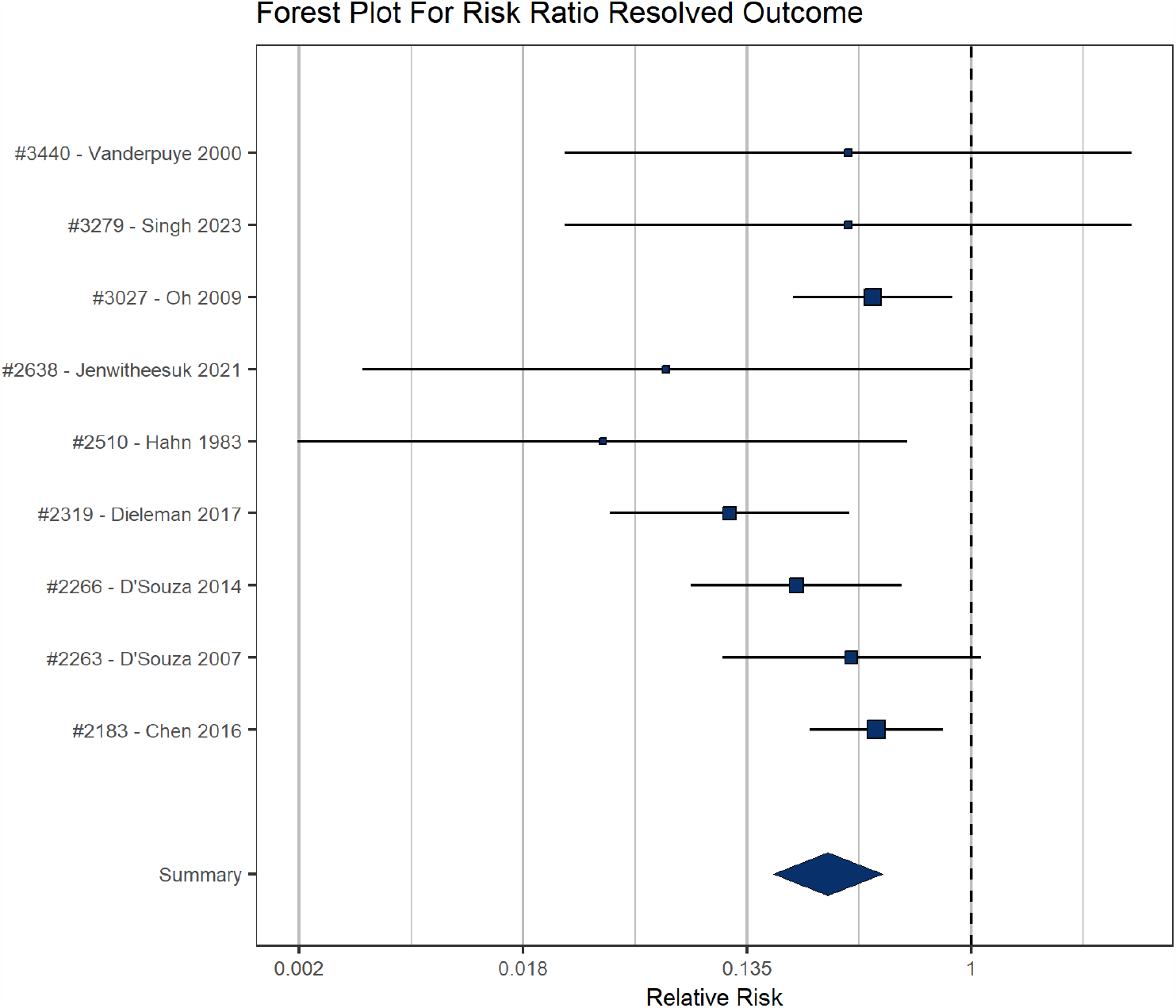
Forest plot demonstrating the risk ratio of resolved outcomes when using HBOT versus surgical free flap.

All nine studies reported on resolution of ORN with HBOT vs. free flap, with six studies showing significant differences in outcomes such that surgical free flap yielded significantly higher complete resolution of ORN compared to HBOT (RR 0.42, 95% CI 0.2-0.85; RR 0.07, 95% CI 0-0.99; RR 0.04, 95% CI 0-0.57; RR 0.12, 95% CI 0.04-0.34; RR 0.21, 95% CI 0.08-0.54; RR 0.43, 95% CI 0.24-0.78). Three studies showed no significant differences in outcomes (RR 0.33, 95% CI 0.03-4.19; RR 0.33, 95% CI 0.03-4.19; RR 0.34, 95% CI 0.11-1.09). The summary outcome for ORN resolution was a significant difference between surgical free flap and HBOT (RR 0.28, 95% CI 0.17-0.45). These results are shown in **Table 3**.

## DISCUSSION

Our findings address outcomes when considering patients who received ORN treatment with curative intent. While two studies included in our meta-analysis address outcomes of improvement as well as resolution, the majority report only on complete resolution of ORN. When comparing rates of ORN resolution when treated with HBOT versus surgical free flap, our study found that those who undergo extensive resection and reconstruction have superior cure rates.

Many studies report that patients only benefit from HBOT when they have less severe disease. When taken as aggregate, the cure rates with HBOT are low, because HBOT has been found to be ineffective in patients with more severe stages of ORN. Considering the low ORN cure rates with HBOT, it is pertinent to consider which patients may benefit from HBOT versus those who require surgical management. Of the studies included in our analysis, almost all patients were stratified using the Marx grading system. Of these patients, patients with stage 1 disease and some patients with stage 2 disease experienced benefit. However, such a grading system requires all patients with ORN to undergo 30 dives of HBOT therapy, which, at 90 minutes per session, is time intensive and costly.

The pathogenesis of ORN was initially hypothesized to involve a triad of radiation, trauma, and infection.^8^ This early definition was challenged by Marx, who noticed that not all cases of ORN involved trauma, and that ORN did not progress to sepsis as seen in other cases of osteomyelitis. He proposed the hypoxic-hypocellular-hypovascular theory, which involved a 4-part process: 1) radiation; 2) formation of hypoxic-hypovascular-hypocellular tissue; 3) tissue breakdown whereby collagen lysis and cellular death exceeds synthesis and cellular replication; and 4) chronic non-healing wounds in which energy, oxygen, and structural precursor demand exceeds supply.^9,10^ Given that the major driver of ORN was proposed to be hypoxia, HBOT emerged as a primary treatment of ORN.^11,10^ HBOT therapy, in theory, helps to increase the oxygen content in hypoxic tissue, stimulating angiogenesis, collagen formation, and fibroblast proliferation. However, while HBOT therapy can work to preserve viable tissue, it has less of a role in healing of tissue that is already nonviable.

A clinically based grading system may help identify patients that would benefit from HBOT therapy before trialing the intervention. For example, Notani et al. proposed a clinical grading system based on mandibular extent of ORN. Grade I was confined to alveolar bone, grade II was limited to the alveolar bone or mandible above the level of the mandibular canal, and grade III extended to mandible under the level of mandibular canal with skin fistula or pathological fracture.^12^ Similar to the Marx grading system, patients with grade I and II disease per Notani et al. experienced good response to conservative management and less extensive surgical excision, supporting the utility of this system in stratifying patients regarding their predicted response to HBOT. Other studies have proposed similar grading systems and stratified treatments by stage of disease.^13,14,15,16^ For future studies, it would be worth analyzing treatment outcomes after separating patients by stage of disease and comparing HBOT versus surgical free flap for patients within each group.

A more recent theory is the radiation-induced fibroatrophic theory, which suggests that the central event causing ORN is the activation and deregulation of fibroblastic activity leading to atrophic tissue within a previously irradiated area. The hypothesis focuses on an imbalance between tissue synthesis and tissue degradation in irradiated tissue. There are 3 distinct phases: 1) the prefibrotic phase, 2) the continuative organized phase, and 3) the late fibroatrophic phase. Endothelial cells undergo injury from radiation as well as from an acute inflammatory response which leads to release of reactive oxygen species. Further release of cytokines including tumor necrosis factor-alpha, platelet-derived growth factor, fibroblast growth factor-beta, interleukin (IL)-1, IL-4, and IL-6, and transforming growth factor-beta-1, result in abnormal fibroblast activity, leading to a disorganized extracellular matrix and healed but fragile tissue that is prone to repeated injury and a local inflammatory response. The resulting bone is hypocellular and fibrotic. As such, more recent treatments focus on targeting reactive oxygen species and the dysregulated fibroblast response.^17,18,10^

PENTOCLO is a recent development including three components, pentoxifylline, tocopherol, and clodronate. Pentoxifylline, a methylxanthine derivative, is thought to help with vascular dilation and improved blood flow to affected tissues. Tocopherol is a vitamin E analog that is thought to play a role in targeting the ROS that promote ORN pathogenesis. Clodronate acts on osteoblasts to increase formation of bone as well as reduce fibroblast proliferation, thereby also inhibiting further ORN pathogenesis based on the radiation-induced fibroatrophic theory.^19,1^ As such, another limitation of the present study is that it does not analyze PENTOCLO’s efficacy as a conservative management option. However, the literature has shown that it yields moderate success in healing ORN. This treatment could be further explored in future studies comparing conservative management to surgical intervention in the treatment of ORN.

Since the present study was conducted to explore the role of HBOT in treating ORN patients with curative intent, a different discussion must take place when considering patients too frail or otherwise uninterested in free flap management. A limitation of the present study was the inability to gather detailed data regarding treatment response, such that it is unknown whether patients with persistent disease experienced worsening, stable, or improved disease. An analysis of such outcomes would facilitate discussions regarding treatment outcomes and expectation management with patients who are ineligible for free flap.

## Data Availability

All data produced in the present work are contained in the manuscript

### APPENDIX

### Methods

Search databases and search terms:

PubMED: Wednesday March 8, 2023

(“osteoradionecrosis”[MeSH] OR “osteoradionecro*” OR “bone radionecro*” OR “osteo-radio-necro*” OR “osteo-radionecro*” OR “radiation bone necros*” OR “radiation osteonecro*” OR “radiation-induced osteonecro*” OR “radiation-induced osteonecro* lesion” OR “radiation-related osteonecro*” OR “radio-osteonecro*” OR “radiotherapy-related osteonecro*”) AND ((“hyperbaric oxygenation”[MeSH] OR “hyperbaric oxygen therapy” OR “HBO-therapy” OR “high pressure oxygen” OR “high tension o2” OR “high tension oxygen” OR “hyperbaric medicine” OR “hyperbaric o2” OR “hyperbaric oxygen” OR “hyperbaric oxygen treatment” OR “hyperbaric oxygenation” OR “hyperbaric oxygenisation” OR “hyperbaric oxygenization” OR “hyperbaric therapy” OR “oxygen, hyperbaric”) OR (“free tissue flaps”[MeSH] OR “free tissue graft” OR “free flap” OR “free flap transfer” OR “free flaps” OR “free microvascular flap” OR “free tissue flap” OR “free tissue flaps” OR “free tissue transfer” OR “graft, free”))

Embase: Wednesday March 8, 2023

(‘osteoradionecrosis’/exp OR ‘osteoradionecro*’ OR ‘bone radionecro*’ OR ‘osteo-radio-necro*’ OR ‘osteo-radionecro*’ OR ‘radiation bone necros*’ OR ‘radiation osteonecro*’ OR ‘radiation-induced osteonecro*’ OR ‘radiation-induced osteonecro* lesion’ OR ‘radiation-related osteonecro*’ OR ‘radio-osteonecro*’ OR ‘radiotherapy-related osteonecro*’) AND ((‘hyperbaric oxygenation’/exp OR ‘hyperbaric oxygen therapy’ OR ‘HBO-therapy’ OR ‘high pressure oxygen’ OR ‘high tension o2’ OR ‘high tension oxygen’ OR ‘hyperbaric medicine’ OR ‘hyperbaric o2’ OR ‘hyperbaric oxygen’ OR ‘hyperbaric oxygen treatment’ OR ‘hyperbaric oxygenation’ OR ‘hyperbaric oxygenisation’ OR ‘hyperbaric oxygenization’ OR ‘hyperbaric therapy’ OR ‘oxygen, hyperbaric’) OR (‘free tissue flaps’/exp OR ‘free tissue graft’ OR ‘free flap’ OR ‘free flap transfer’ OR ‘free flaps’ OR ‘free microvascular flap’ OR ‘free tissue flap’ OR ‘free tissue flaps’ OR ‘free tissue transfer’ OR ‘graft, free’))

Clinicaltrials.gov March 8, 2023

Condition or disease: “Osteoradionecrosis of Jaw”

Other terms: hyperbaric oxygen therapy OR free flap

#### Inclusion criteria

- Studies that analyze the healing outcomes from interventions including, but not limited to, hyperbaric oxygen therapy AND/OR free tissue flap/graft in the management of osteoradionecrosis of the jaw secondary to radiation therapy (curative or adjunctive)
- Studies conducted on head and neck cancer patients diagnosed with osteoradionecrosis of the jaw, based on clinical or imaging characteristics
- Study design types to be included: randomized controlled trials, prospective and retrospective cohort studies, case studies, case reports
- Studies investigating osteoradionecrosis of the jaw secondary to radiation therapy for non-head and neck cancer
- No language restriction

#### Exclusion criteria

- Studies in which there is no or inadequate follow-up (less than 1 year)
- Studies assessing interventions that do not include HBOT or free tissue flap/graft for management of osteoradionecrosis of the jaw
- Studies assessing treatment outcomes of osteoradionecrosis of the jaw as a secondary outcome
- Studies analyzing ORN caused by interventions other than radiation therapy (bisphosphonates, denosumab, etc.)
- Studies that are observational, including systematic reviews
- Patients treated by radiation therapy with palliative intent, due to differences in treatment goals, increased incidence of extant comorbidities, and lack of universally-accepted standardized palliative radiotherapy regimens, most of which involve a total radiation dose of <u>less than 60 Gy</u> with <u>minimal associated risk for development of ORN</u>, as well as critically-reduced overall survival rates leading to severely constrained follow-up times
- Re-irradiated patients due to the permanent/irreversible effects associated with prior radiation treatment which would result in inextricable confounding
- Child and adolescent populations (age <18) undergoing radiotherapy are unique and present with different concerns due to the distinct biological behavior of developing bone, and should thus be considered in a separate review (and excluded from this study)
- Complex infections of the mandible

## TABLES

**Table 1.** Summary of final included studies.

| Study | Title | Country | Study design | Total number of participants |
| --- | --- | --- | --- | --- |
| Vanderpuye 2000 | Osteoradionecrosis of the mandible | United States | Case report | 1 |
| Singh 2023 | Osteoradionecrosis of the Jaw Following Proton Radiation Therapy for Patients With Head and Neck Cancer | United States | Cohort study | 2 |
| Oh 2009 | Osteoradionecrosis of the mandible: treatment outcomes and factors influencing the progress of osteoradionecrosis | South Korea | Cohort study | 37 |
| Jenwitheesuk 2021 | Is Adjunctive Hyperbaric Oxygen Treatment Alone or with Surgery the Proper Management for Active and Persistent Osteoradionecrosis? | Thailand | Case control | 61 |
| Hahn 1983 | Osteoradionecrosis of the mandible: clinical observation and treatment in 45 cases | Taiwan | Cohort study | 45 |
| Dieleman 2017 | The efficacy of hyperbaric oxygen therapy related to the clinical stage of osteoradionecrosis of the mandible | Netherlands | Cohort study | 27 |
| D'Souza 2014 | Changing trends and the role of medical management on the outcome of patients treated for osteoradionecrosis of the mandible: experience from a regional head and neck unit | UK | Cohort study | 41 |
| D'Souza 2007 | The influence of hyperbaric oxygen on the outcome of patients treated for osteoradionecrosis: 8 year study | UK | Cohort study | 20 |
| Chen 2016 | Osteoradionecrosis of mandible bone in patients with oral cancer - Associated factors and treatment outcomes | Taiwan | Cohort study | 105 |

**Table 2.** Risk-ratio calculation for ORN improvement outcomes.

| v4 | RiskRatio |  |  |
| --- | --- | --- | --- |
| #2319 - Dieleman 2017 | RR: 0.2; CI 0.09, 0.42 |  |  |
| #2266 - D'Souza 2014 | RR: 0.33; CI 0.07, 1.51 |  |  |
| v4 | Risk Ratio | I2 | H2 |
| Summary Outcome | RR: 0.22; CI 0.02, 2.94 | 0 | 1 |

**Table 3.** Risk-ratio calculation for ORN resolution outcomes.

| v4 | RiskRatio |  |  |
| --- | --- | --- | --- |
| #3440 - Vanderpuye 2000 | RR: 0.33; CI 0.03, 4.19 |  |  |
| #3279 - Singh 2023 | RR: 0.33; CI 0.03, 4.19 |  |  |
| #3027 - Oh 2009 | RR: 0.42; CI 0.2, 0.85 |  |  |
| #2638 - Jenwitheesuk 2021 | RR: 0.07; CI 0, 0.99 |  |  |
| #2510 - Hahn 1983 | RR: 0.04; CI 0, 0.57 |  |  |
| #2319 - Dieleman 2017 | RR: 0.12; CI 0.04, 0.34 |  |  |
| #2266 - D'Souza 2014 | RR: 0.21; CI 0.08, 0.54 |  |  |
| #2263 - D'Souza 2007 | RR: 0.34; CI 0.11, 1.09 |  |  |
| #2183 - Chen 2016 | RR: 0.43; CI 0.24, 0.78 |  |  |
| v4 | Risk Ratio | I2 | H2 |
| Summary Outcome | RR: 0.28; CI 0.17, 0.45 | 21.8 | 1.28 |

## FIGURES

**allFigure 1.**
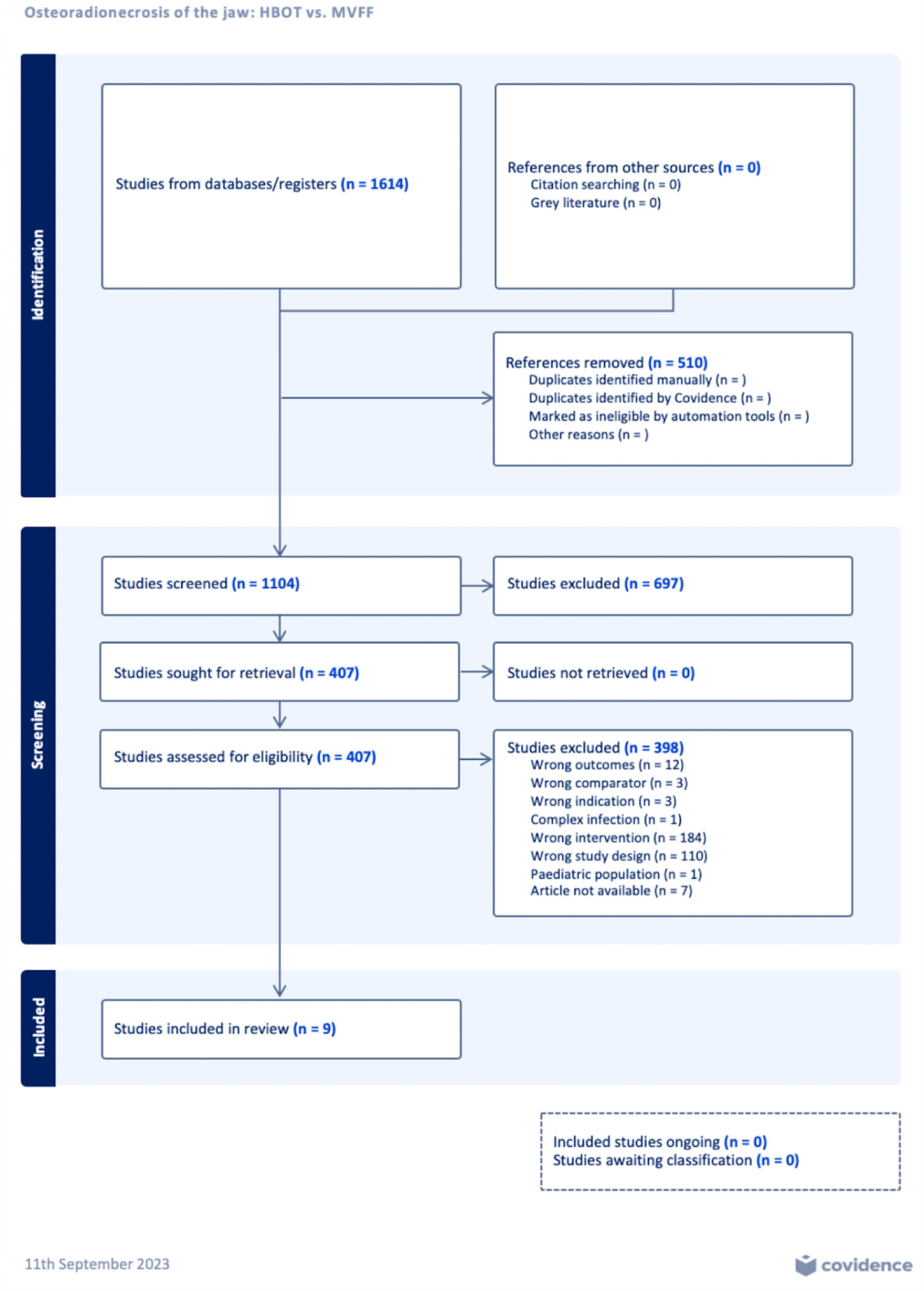
PRISMA diagram.

## REFERENCES

1. Vahidi N, Lee TS, Daggumati S, Shokri T, Wang W, Ducic Y. Osteoradionecrosis of the Midface and Mandible: Pathogenesis and Management. Semin Plast Surg. 2020;34(4):232–244. doi:10.1055/s-0040-1721759

2. Moon DH, Moon SH, Wang K, et al. Incidence of, and risk factors for, mandibular osteoradionecrosis in patients with oral cavity and oropharynx cancers. Oral Oncol. 2017;72:98–103. doi:10.1016/j.oraloncology.2017.07.014

3. Chen JA, Wang CC, Wong YK, et al. Osteoradionecrosis of mandible bone in patients with oral cancer--associated factors and treatment outcomes. Head Neck. 2016;38(5):762–768. doi:10.1002/hed.23949

4. Balermpas P, van Timmeren JE, Knierim DJ, Guckenberger M, Ciernik IF. Dental extraction, intensity-modulated radiotherapy of head and neck cancer, and osteoradionecrosis : A systematic review and meta-analysis. Strahlenther Onkol. 2022;198(3):219–228. doi:10.1007/s00066-021-01896-w

5. Rice N, Polyzois I, Ekanayake K, Omer O, Stassen LFA. The management of osteoradionecrosis of the jaws--a review. Surgeon. 2015;13(2):101–109. doi:10.1016/j.surge.2014.07.003

6. Marx RE, Ames JR. The use of hyperbaric oxygen therapy in bony reconstruction of the irradiated and tissue-deficient patient. J Oral Maxillofac Surg. 1982;40(7):412–420. doi:10.1016/0278-2391(82)90076-3

7. Gevorgyan A, Wong K, Poon I, Blanas N, Enepekides DJ, Higgins KM. Osteoradionecrosis of the mandible: a case series at a single institution. J Otolaryngol Head Neck Surg. 2013;42(1):46. doi:10.1186/1916-0216-42-46

8. Meyer I. Infectious diseases of the jaws. J Oral Surg. 1970;28(1):17–26.

9. Marx RE. Osteoradionecrosis: a new concept of its pathophysiology. J Oral Maxillofac Surg. 1983;41(5):283–288. doi:10.1016/0278-2391(83)90294-x

10. O’Dell K, Sinha U. Osteoradionecrosis. Oral Maxillofac Surg Clin North Am. 2011;23(3):455–464. doi:10.1016/j.coms.2011.04.011

11. Marx RE. A new concept in the treatment of osteoradionecrosis. J Oral Maxillofac Surg. 1983;41(6):351–357. doi:10.1016/s0278-2391(83)80005-6

12. Notani KI, Yamazaki Y, Kitada H, et al. Management of mandibular osteoradionecrosis corresponding to the severity of osteoradionecrosis and the method of radiotherapy. Head Neck. 2003;25(3):181–186. doi:10.1002/hed.10171

13. Kün-Darbois JD, Fauvel F. Medication-related osteonecrosis and osteoradionecrosis of the jaws: Update and current management. Morphologie. 2021;105(349):170–187. doi:10.1016/j.morpho.2020.11.008

14. Glanzmann C, Grätz KW. Radionecrosis of the mandibula: a retrospective analysis of the incidence and risk factors. Radiother Oncol. 1995;36(2):94–100. doi:10.1016/0167-8140(95)01583-3

15. Støre G, Boysen M. Mandibular osteoradionecrosis: clinical behaviour and diagnostic aspects. Clin Otolaryngol Allied Sci. 2000;25(5):378–384. doi:10.1046/j.1365-2273.2000.00367.x

16. Schwartz HC, Kagan AR. Osteoradionecrosis of the mandible: scientific basis for clinical staging. Am J Clin Oncol. 2002;25(2):168–171. doi:10.1097/00000421-200204000-00013

17. Delanian S, Lefaix JL. The radiation-induced fibroatrophic process: therapeutic perspective via the antioxidant pathway. Radiother Oncol. 2004;73(2):119–131. doi:10.1016/j.radonc.2004.08.021

18. Delanian S, Depondt J, Lefaix JL. Major healing of refractory mandible osteoradionecrosis after treatment combining pentoxifylline and tocopherol: a phase II trial. Head Neck. 2005;27(2):114–123. doi:10.1002/hed.20121

19. Rivero JA, Shamji O, Kolokythas A. Osteoradionecrosis: a review of pathophysiology, prevention and pharmacologic management using pentoxifylline, α-tocopherol, and clodronate. Oral Surg Oral Med Oral Pathol Oral Radiol. 2017;124(5):464–471. doi:10.1016/j.oooo.2017.08.004

